# Predicting ovarian/breast cancer pathogenic risks of BRCA1 gene variants of unknown significance

**DOI:** 10.1101/2020.06.04.20120055

**Authors:** Hui-Heng Lin, Hongyan Xu, Hongbo Hu, Zhanzhong Ma, Jie Zhou, Qingyun Liang

## Abstract

The difficulty of early diagnosis for ovarian cancer is an important cause of the high mortal rates of ovarian cancer patients. Instead of symptom-based diagnostic methods, modern sequencing technologies enable the access of human’s genetic information via reading DNA/RNA molecules’ nucleotide base sequences. In such way, genes’ mutations and variants could be identified and hence a better clinical diagnosis in molecular level could be expected. However, as sequencing technologies gain more popularity, novel gene variants with unknown clinical significance are found, giving difficulties to interpretations of patients’ genetic data, precise disease diagnoses as well as the making of therapeutic strategies and decisions. In order to solve these issues, it is of critical importance to figure out ways to analyze and interpret such variants. In this work, BRCA1 gene variants with unknown clinical significance were identified from clinical sequencing data, and then we developed machine learning models so as to predict the pathogenicity for variants with unknown clinical significance. Amongst, in performance benchmarking, our optimized random forest model scored 0.85 in area under receiver-operating characteristic curve, which outperformed other models. Finally, we applied the optimized random forest model to predict the pathogenic risks of 7 BRCA1 variants of unknown clinical significances identified from our sequencing data, and 6315 variants of unknown clinical significance in ClinVar database. As a result, our model predicted 4724 benign and 1591 pathogenic variants, which helped the interpretation of these variants of unknown significance and diagnosis.

## Introduction

For diagnosis of ovarian cancer, the symptom-based diagnostic approaches tend to be less precise because it usually do not display obvious and specific symptoms on early stage patients. Therefore, unfortunately, the usual cases are that, when confirmed, the cancer are already developed to a late stage. The difficulties in detecting specific symptoms in early stage ovarian cancer have affected precise diagnosis and it is one of the important reasons causing the high mortal rate of ovarian cancer [1] [2] [3].

Since ovarian cancer is a multi-genic disease, in addition that limitations exist in symptom-based diagnostic methods, molecular genetic diagnosis could be a better way for ovarian cancer diagnosis, especially in cases of early stage. According to investigations, several genes are associated with the pathogenesis of ovarian cancer, and amongst, two genes — the BRCA1 and BRCA2, are well known ones and found to have significant associations with ovarian cancer [4]. “BRCA1” standards for “BReast CAncer type 1 protein”. It is a tumor suppressor and found to be associated with familial breast cancer [5]. Since the discovery of BRCA1, scientists keep researching on the molecular structure and functions of it and its products [6] [7] [8] [9] Thanks to their elaborated efforts and works, parts of its functions and roles in biological processes have been elucidated. For instance, BRCA1 is known to participate the processes of DNA repairing [10] [11], and its mutations/variants are known to have association with the onset of breast cancer and ovarian cancer [12] [13] [14]. Moreover, germline mutations of BRCA1 have also been discovered, and it is reported that, for patients of familial ovarian cancer, over 80% patients carry BRCA1 (or BRCA2) mutation [4] [15] [16]. Because of the high susceptibility of BRCA1 to ovarian cancer, for molecular and genetic tests of ovarian cancer, BRCA1 is one of the most indispensable genes for probing.

With advancements of sequencing technologies, besides research laboratories, nowadays high-throughput sequencers are also being employed in clinical contexts. Especially for clinical diagnoses in genetic and molecular level. The state-of-art sequencing technology enables the detection and reading of nucleotide sequences in human body, which greatly facilitates the clinical diagnosis in terms of molecular pathology and medicine. Though sequencing technologies are acquiring popularity from clinical diagnoses, several challenges are encountered. One of the most significant challenges is the interpretation of gene Variants of Unknown Significance (VUSs). Sequencing real-world human tissue samples has been generating large sets of novel gene variant data. However, except their physical/structural variation information, such as the mutation position, and nucleic acids’ changes, nothing else is known. This gives great difficulties in data interpreting and clinical diagnoses. For example, real cases happened that, novel VUSs of BRCA1 were detected in multiple people who looked like healthy (at least no symptom was detected), while doctors still had no way to conclude the pathogenic risks for these people since nothing but just the physical variant information. Therefore, it is urgent and necessary to find ways to interpret VUSs of genes.

Theoretically, biomedical characteristics and molecular functions of a VUS can be explored via biochemical approached. E.g., immunocoprecipitation and immunoblotting can probe the potential molecules that have binding interactions with the protein of VUSs. Crystallization and 3D structural visualization approaches can help reveal the structure-function relationships of the protein of VUSs. While these methods are highly costly in all sorts of aspects including time, labor forces, budget, etc. In fact, large amounts of such VUSs are seen in clinical data and doctors usually face more than one VUSs. Thus, for the purpose of quickly and efficiently characterizing large amounts of VUSs, wet lab methods are basically impractical. The more realistic and practical way is computational analysis.

In present study, BRCA1 VUSs were identified from clinical sequencing data. (Note that in this study, we also consider “likely pathogenic” and “likely benign” variants as the VUSs, since these variants lack solid evidence that can demonstrate their pathogenic risks). In order to interpret the clinical significances of these data, we analyzed the data and developed multiple machine learning models to predict the pathogenic risks of these VUSs. After benchmarking, the optimized random forest model was found to have the best performance and it was chosen to predict BRCA1 VUSs from both our sequencing data and ClinVar database [17]. As a result, predictive pathogenicity of total 6322 VUSs were obtained. Amongst, 1593 VUSs were predicted to be pathogenic and 4729 VUSs were predicted to be benign.

## Results

### BRCA1 VUS data

Through sequencing data analyses and database queries, we totally identified 7 BRCA1 VUSs that have substitution of single nucleotide base. The relevant data and information are shown in **Table 1**. Of 7 BRCA1 variants listed in **Table 1**, 4 of which have unknown clinical significance. Here the term “unknown clinical significance” indicates that, whether or not these VUSs will increase the risk of having relevant diseases remains unclear. For the rest of other 3 BRCA1 variants, they are annotated as “likely benign” in databases. This indicates that so far these variants lack sufficient or solid evidences supporting their associations with pathogenic risk levels, and these uncertainties give difficulties to diagnosis, as well. Hence, the word “likely” is used and it is necessary to further analyze these variants. Specifically, one of our 7 VUSs, the “c.1348A>T”, could not be found in results of database queries or searching engines. It is indicating that, the BRCA1 “c.1348A>T” variant is a new variant identified from our sequencing data. Regarding this novel BRCA1 variant and its molecular functions, though nothing is known except its sequence variation information, through our computational analyses and machine learning prediction, its pathogenicity was characterized.

**Table 1.** 7 VUSs of BRCA1 identified from sequencing data and databases.

| ID | Variation | refSNP ID (a.k.a. Rs number) | Clinical significance in database |
| --- | --- | --- | --- |
| 1 | c.1255G>C | rs876658873 | Unknown |
| 2 | c.824G>A | rs397509327 | Unknown |
| 3 | c.3448C>T | rs80357272 | Unknown |
| 4 * | c.1348A>T | Not available | Unknown |
| 5 | c.2566T>C | rs80356892 | Likely benign |
| 6 | c.3748G>A | rs28897686 | Likely benign |
| 7 | c.571G>A | rs80357090 | Likely benign |

\* This BRCA1 variant was not found through querying databases.

### Performances of predictive models

As described in methods section, initially 5 predictors were chosen for present study. While because of the obviously biased performance of naïve bayes in primary tests [18], it was excluded from further analysis and only 4 other predictors were used. Upon benchmarking with datasets, each model’s receiver-operating characteristic curve (ROC) was plotted and the relevant value of area under curve (AUC) was computed accordingly. Eventually, the performances of 4 included models are as shown in **Figure 1**.

**Figure 1.**
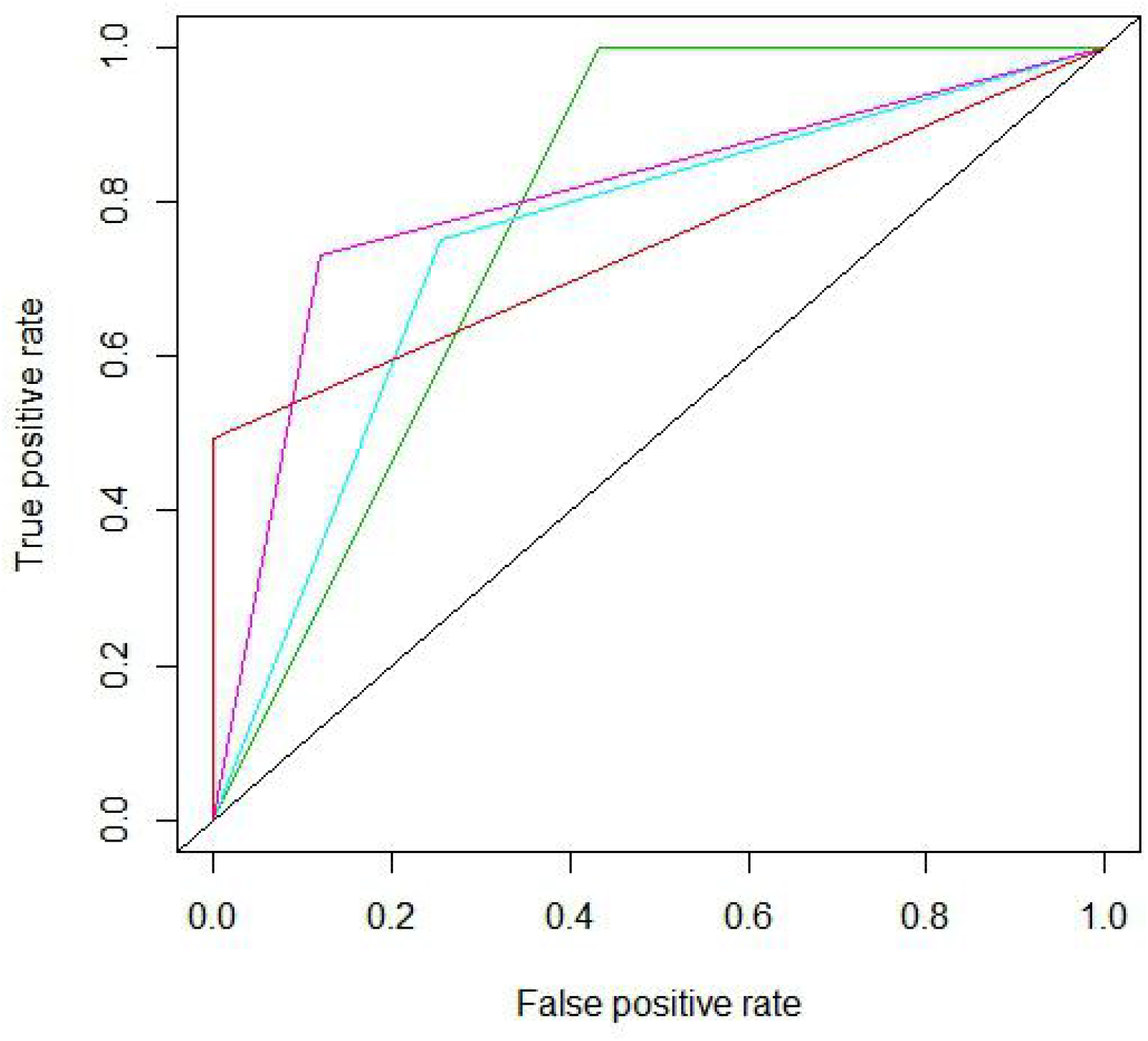
ROCs of 4 kinds of predict models. ROCs indicating varied performances of different models were plotted. And the relevant AUCs were also computed to indicate models’ overall performance. For support vector machine (the light blue curve), random forest (the purple curve), Polyphen (the red curve), and SIFT (the green curve), their AUC values were 0.74, 0.78, 0. 74 and 0.78, respectively.

The 4 tested models, i.e., the support vector machine [19], the random forest [20], the Polyphen [21] [22], and the SIFT [23], have the AUC values of 0.74, 0.78, 0.74, and 0.78, respectively. For our own models, the random forest outperformed the support vector machine. For comparison between our own models and others’ models, the support vector machine had similar overall performance with Polyphen, and the random forest had similar performance with SIFT as well.

Since our own models had similar performances with the two reference models developed by other researchers, in order to obtain the better model, we subsequently tried to optimize our own support vector machine and random forest models, hoping to see improvements in our models’ predictive performances. The optimization works are described in the methods section and the performances of optimized models are as seen in **Figure 2**.

**Figure 2.**
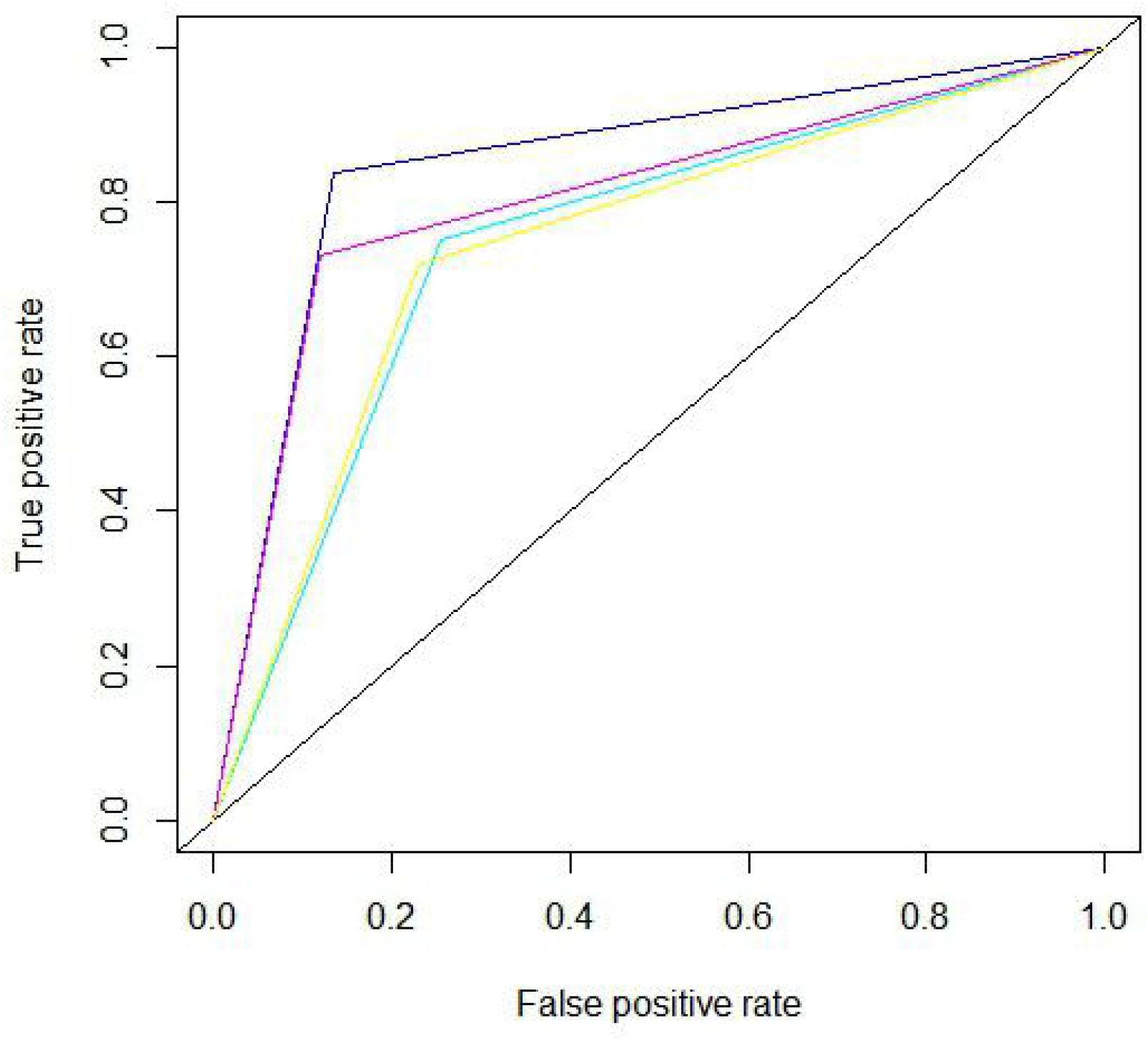
The overall performance of optimized support vector machine, optimized random forest model, original (not optimized) support vector machine and original random forest. The optimized random forest (dark blue) had obvious larger AUC than the optimized support vector machine (light blue) and the original random forest (purple). While no significant increase of AUC was observed between the original support vector machine (yellow) and the optimized one (light blue). The quantified AUC value of optimized random forest and optimized support vector are 0.85 and 0.75, respectively, indicating that the random forest model had better performance after optimization, while support vector machine did not.

For both optimized models, the AUC of ROC indicates that the optimized random forest model had better overall performance than the optimized support vector machine. While comparing with our own models developed before optimization works, support vector machine seemed to have little improvement (merely very slight improvement from 0.74 to 0.75, only 0.01’s difference in AUC value). And for the random forest model, the optimization helped random forest model improve its AUC value from 0.78 to 0.853 (the best values). Specifically, in order to detect the robustness of model’s performance, we further carried out 10 times of 10-fold cross validation to test the optimized random forest model. As a result, the mean value and standard deviation of ACUs are 0.85 and 0.006, respectively. This result showed that the optimized random forest had stable performance. For the best model of random forest, in **Table 2**, we listed its other performance indicators based on confusion matrix computing. Overall, the optimized random forest model had good performance.

**Table 2.** Other performance indicators of the best random forest model. Tp, Tn, Fp and Fn stand for the number of true positive, true negative, false positive and false negative instance in the machine learning confusion matrix, respectively

| ID | Indicator | Value | Calculation |
| --- | --- | --- | --- |
| 1 | True positive rate (a.k.a Sensitivity or Recall) | 0.84 | $Tp / (Tp + Fn)$ |
| 2 | True negative rate (a.k.a specificity) | 0.86 | $Tn / (Tn + Fp)$ |
| 3 | False positive rate | 0.13 | $Fp / (Fp + Tn)$ |
| 4 | False negative rate | 0.16 | $Fn / (Fn + Tp)$ |
| 5 | Positive predictive value (a.k.a Precision) | 0.77 | $Tp / (Tp + Fp)$ |
| 6 | Accuracy | 0.85 | $(Tp + Tn) / (Tp + Tn + Fp + Fn)$ |
| 7 | Balanced accuracy | 0.85 | $(\text{True positive rate} + \text{True negative rate}) / 2$ |
| 8 | F-measure (a.k.a F1-score) | 0.80 | $2Tp / (2Tp + Fp + Fn)$ |

### Prediction of pathogenic risks of BRAC1 VUSs

Upon identification of the model with best predictive performance, i.e., the optimized random forest, it has been applied to predict the BRCA1 VUSs’ pathogenic risks for ovarian/breast cancer. We initially predicted the BRCA1 VUSs identified from our sequencing dataset. The results of the machine learning predictive analysis are shown in **Table 3**.

**Table 3.** Predictive pathogenic risks for 7 VUSs of BRCA1 identified from our sequencing data.

| ID | Variation | (Original) Clinical<br>significance | Predictive pathogenic<br>risk |
| --- | --- | --- | --- |
| 1 | c.1255G>C | Unknown | Pathogenic |
| 2 | c.824G>A | Unknown | Benign |
| 3 | c.3448C>T | Unknown | Benign |
| 4 * | c.1348A>T | Unknown | Pathogenic |
| 5 | c.2566T>C | Likely benign | Benign |
| 6 | c.3748G>A | Likely benign | Benign |
| 7 | c.571G>A | Likely benign | Benign |

\* Except this variant, the rest variants can be found in ClinVar database

For the first 3 VUSs, our best model predicted the first VUS to be pathogenic while the other 2 are benign (**Table 3)**. And for the rest 4 VUSs, according to our predictive analysis, their prefix word “likely” was removed from their original clinical significances. In the other word, our model had given further confidence to the likelihood of database’s original pathogenic annotations of them.

Additionally, we found that large amounts of VUSs exist in ClinVar database, too. Many gene variants were discovered and submitted to ClinVar. However, as mentioned before, parts of variants do not have any function annotation and hence they remain to be the VUSs (Note that, here for variants in ClinVar, we consider those variants of “likely benign”, “likely pathogenic”, “not provided” and “conflicting interpretations of pathogenicity” the same as those variants of “uncertain significance”, as the VUSs. And hence all of them were the targets for our predictive analysis). For example (as of 16 May 2020), for BRCA1, the number of variants of “likely benign”, “likely pathogenic”, “not provided”, “conflicting interpretations of pathogenicity” and “uncertain significance” are 937, 82, 2797, 264, and 2235, respectively (**Table 4**). In order to provide more insights for these BRCA1 VUSs which are not covered by our datasets, we also used the optimized random forest model to carry out pathogenic risk prediction these BRCA1 VUSs. Notice that, 6 of VUSs identified from sequencing data were also found in ClinVar’s VUSs. For total 6315 predicted variants, in which 6 VUSs identified from sequencing were excluded, 4724 (74.81%) were predicted to be benign and 1591 (25.19%) were predicted to be pathogenic (**Table 4**). Interestingly, we found that the percentages of predicted pathogenic variants in different subclasses are close. These percentages range from 69.51% to 77.80%. For variants of “Likely pathogenic”, “Uncertain significance”, and “Conflicting interpretations of pathogenicity”, the predicted pathogenic ratios of variants are 69.51%, 71.19% and 71.97%, respectively. These values fluctuate around 70%. While the rest two percentages, 77.80% for “Not provided” subgroup of BRCA1 VUSs and 75.78% for “Likely benign”, are slightly higher than the former 3 percentages **(Table 4)**. The full predictive results can be found in **supplementary file S1**.

**Table 4.** Overview of predictive results for 6321 BRCA1 VUSs pathogenic risks

| **Class** | **Clinical significance** | **Variant amount** | **Number of** | **Number of** |
| --- | --- | --- | --- | --- |
|  | **(ClinVar database)** |  | **predictive** | **predictive** |

|  |  |  | **pathogenic** | **benign** |
| --- | --- | --- | --- | --- |
|  |  |  | **variants (%)** | **variants (%)** |
| 1 | Likely benign | 937 | 227 (24.22%) | 710 (75.78%) |
| 2 | Likely pathogenic | 82 | 25 (30.49%) | 57 (69.51%) |
| 3 | Not provided | 2797 | 621 (22.20%) | 2176 (77.80%) |
| 4 | Uncertain significance | 2235 | 644 (28.81%) | 1591 (71.19%) |
| 5 | Conflicting interpretations<br>of pathogenicity | 264 | 74 (28.03%) | 190 (71.97%) |
| 6 | Total | 6315 | 1591 (25.19%) | 4724 (74.81%) |

## Methods

### Datasets and processing

Initially, we identified and processed BRCA1 variant data from RNA-seq. Since year 2017, we have started to ask hospital visitors that if they are willing to donate their samples for our cancer genetic research purposes. For those who agreed, we sampled their bloods, and made a third-party contractor research organization (Beijing Genome Institute, Shenzhen, China) to perform RNA-seq to these samples. After bioinformatic analyses, we further confirmed, analyzed and identified the BRCA1 variant data via queries against databases (Ensembl [24], dbSNP [25], and ClinVar [17], as of 15 April 2020), information parsing and data cleaning. Accordingly these variation data were parsed and converted into DNA sequences and they were prepared for loading into predictive model predicting risks of ovarian/breast cancer

Next, we prepared the dataset for training and benchmarking predictive models. We extracted BRCA1’s variant data from the ClinVar, dbSNP, and Ensembl databases following such criteria: (1) Only retrieve BRCA1 variant data that are labeled “benign” and “pathogenic” for ovarian/breast cancer. (2) Choose variants that only have single nucleotide base substitution. (3) Choose variants that are reviewed by expert panel. As a result, 499 pathogenic BRCA1 variants and 585 benign BRCA1 variants were obtained. These variant data were further transformed into DNA sequences, and subsequently, through different type of molecular descriptors, DNA sequences were converted into feature vectors. Totally, more than 100 kinds of feature combinations were tested. Lastly, we found that the combination of 117 features gave the best performance for machine learnings. The adopted feature set included vectors of DNA 3-mer [26], genomic location of variants, and di- (tri-) nucleotide-based auto-cross covariance [27] [28].

### Predictors and performance benchmarking

In order to obtain good predictive results, we selected multiple predictors, and benchmarked their performance. Through benchmarking, we chose the most well performed model to predict the pathogenic risk of BRCA1 VUSs so as to obtain the better and more precise results.

After preparation of datasets, we initially chose 5 predictors, i.e., the naïve bayes [18], the support vector machine [19], the random forest [20], the polyphen program [22], and the SIFT program [23]. Amongst, the former 3 are classic machine learning models, and we trained, generated and tested the models by ourselves. While for the latter 2 were programs developed by other researchers, their performances were used as the references to our models’. A primary performance test (used 8:2 ratio to randomly split dataset into training set and testing set) on 5 predictors showed that, the naïve bayes had strongly biased results (data were not shown) on our datasets, thus naïve bayes was excluded from further analyses.

A series of methods were used for machine learning model optimizations. E.g., we adopted oversampling method for balancing the positive and negative training set. We applied 10-fold cross validation strategy to models. We tried to select feature set giving the best performance to models. We also tried to perform standardization, normalization, principal component analysis, etc., to our dataset. Specially, for random forest and support vector machine, model-specific optimizations were conducted. For support vector machine, the cost coefficient, gamma, kernel, etc., were tuned. For random forest, specific parameter tunings including the number of trees and number of features, and were carried out.

Predictive performances of all predictors were visualized as ROC plot, and the values of AUC were calculated for quantitative comparison. Accordingly, we also computed optimized random forest’s true positive rate, true negative rate, false positive rate, false negative rate, positive predictive values, accuracy, balanced accuracy, f-measure, so as to examine its extra performance scores in different perspectives.

### Prediction of VUSs

Upon identification of the machine learning model with the best performance in benchmarking. Both sets of BRCA1 VUSs, including 6 VUSs identified from sequencing data and 6315 VUSs identified from ClinVar database, were loaded into the optimized random forest for prediction. The obtained predictive results were statistically analyzed and then compared with their original pathogenicity annotations in the database.

Aforementioned data processing and computational analytic tasks including data parsing, data cleaning, sequence manipulations, format conversions, statistical analyses, numeric computations, etc., were done in R computing environment and Rstudio [29] [30]. Besides self-scripted analytic pipelines, other used R packages include BioMedR [31], Bioconductor [32], Biostring [33], e1071 [34], ROCR [34], RandomForest [20].

## Conclusions

In this work, we identified BRCA1 VUSs from sequencing data, and subsequently we developed machine learning predictive models and benchmarked the performance of predictive models. The best predictive model scored AUC value 0.85, namely the optimized random forest, was used to predict the pathogenic risk of BRCA1 VUSs, including those in the ClinVar database. In total, 6322 variants of unknown clinical significance were predicted. Amongst, one variant “c.1348A>T” identified from the sequencing data has not been found in databases, and hence we considered as a novel VUS. And it was predicted to be pathogenic. For other 6 VUSs identified from sequencing data, “c.1255G>C” was predicted to be pathogenic, as well, while the rest 5 were predicted to be benign. For VUSs in ClinVar, our model predicted 4724 benign variants and 1591 pathogenic ones.

We believe the present study of us is of high significance. Through analyzing potential pathogenicity of BRCA1 VUSs via machine learning approaches, we succeeded in carrying out such translational research that can help clinicians interpret the clinical significances of VUSs. Our work not only facilitates the precise clinical diagnosis, but also provide references to clinical therapeutic decision making.

## Discussion

Before we had the performance scores of the optimized machine learning models, we expected the AUC value of our best model could reach around 0.9. However it did not despite our efforts and multiple trials. While we believe there exist ways for improvement of predictive model, such as using other advanced machine learning models or other optimization methods. For example, through feature engineering approaches, more advance feature sets could be generated and tested.

In present work, our analyses only covered single base substitutions of BRCA1 VUSs. While the forms of BRCA1 VUSs are diverse. Besides single base substitution mentioned in this work, there are deletion, insertion, as well as other kinds of structural variations as well. These variations and variants, though are more complex than single nucleotide base substitution, are of equally high importance for medical researches, clinical data interpretation and clinical diagnosis. In the future, we may take this challenge and try to analyze these more complex forms of BRCA1’s VUSs, so as to acquire more insights between BRCA1 VUSs and cancers.

Variants’ precise functional annotations and associated information are prerequisite for gene-disease relationship analysis and thus play vital and indispensable role in precise diagnosis. On one hand, we know that, the precise functional annotations of gene variants come from the labor-, time- and resource-consuming biochemical assays. On the other hand, more and more novel VUSs are being discovered. And the discovery of VUSs are in more faster speed than the carryout speed of biochemical experiments for VUSs characterization. But for the purposes of clinical diagnosis, no doubt that it is necessary to efficiently interpret the VUSs. Hence, before databases can accumulate and disclose sufficient biochemical assay-based annotation data for gene variants, computational methods for variant analysis would still play important roles in the long run.

## Data Availability

For BRCA1 gene VUS data, they are accessible and stored in NCBI ClinVar database.
For the sequencing data, they will be available upon reasonable requests.

https://www.ncbi.nlm.nih.gov/clinvar/

## Acknowledgements

Not available

